# Early-Onset Complete Atrioventricular Block – Prevalence, Etiology and Utilization of Cardiac Implantable Electronic Devices

**DOI:** 10.1101/2023.03.27.23287825

**Authors:** Gilad Margolis, Jennifer Chee, Mark Kazatsker, Ariel Roguin, Christopher Madias, Munther Homoud, Ofer Kobo, Nashed Hamuda, Inon Dimri, E.Kevin Heist, Jeremy N. Ruskin, Eran Leshem, Guy Rozen

## Abstract

**Background:** Information regarding the prevalence and etiologies of complete atrioventricular block (CAVB) in younger patients is scarce. We aimed to investigate the potential causes for non-iatrogenic CAVB, the prevalence of CAVB without an identified etiology, the utilization of guidelines-recommended advanced imaging modalities in adults presenting with an early-onset CAVB of unidentified etiology, as well as to identify the predictors for cardiac implantable electronic device (CIED) insertion.

**Methods:** Using the National Inpatient Sample (NIS) database, we identified patients aged 18-60 hospitalized with non-iatrogenic CAVB in the US between 2015 (last quarter)-2019. Baseline demographics, clinical characteristics, potential etiologies for CAVB, advanced imaging utilization as well as outcomes including the need for temporary cardiac pacing (TCP) and CIED implantation were analyzed. Multivariable logistic regression models were used to identify predictors of CIED implantation.

**Results:** An estimated total of 14,495 patients aged 18-60 with non-iatrogenic CAVB were identified. The mean age was 51 years, 60% were males and 3,050 (21%) had documentation of a prior conduction disorder. Eleven percent of the patients had a diagnosis of syncope and 6% suffered from a cardiac arrest. Two third of the patients (9,735, 67%) had no identified etiology for CAVB, of whom 8,205 (84%) were implanted with a permanent pacemaker (PPM), 180 patients (2%) with an implantable cardioverter-defibrillator (ICD), and 295 patients (3%) with a cardiac resynchronization therapy device. Only 40 patients (0.3%) underwent advanced imaging during their hospitalization. In multivariate analyses, older age [adjusted OR 1.046 (1.04-1.05), p<0.001] and the need for TCP [adjusted OR 1.543 (1.29-1.84), p<0.001], emerged as predictors for PPM implantation. Cardiac arrest [adjusted OR 2.786 (1.69-4.58), p<0.001] and younger age [adjusted OR 0.98 (0.96-0.99), p=0.02], were associated with ICD implantation. 185 patients (1.3%) died during their hospitalization.

**Conclusion:** The majority of patients, hospitalized in the US for non-iatrogenic early-onset CAVB, had no identified etiology for their conduction disease. Despite the current US and European guidelines recommendation, advanced imaging prior to CIED implantation was under-utilized in this patient population.

## INTRODUCTION

High degree atrioventricular block (AVB) is associated with poor survival rate when managed conservatively (i.e. non-paced)^1^, and with a survival rate comparable to the general population after permanent pacemaker (PPM) implantation in patients without concomitant cardiac disease.^2^ AVB affects most commonly older patients in which ageing-related idiopathic calcification and fibrosis of the conduction system is thought to be the most prevalent etiology (Lev-Lenegre disease).^3, 4^ However, in younger patients, other disease processes may underlie the conduction abnormalities. These include, among others, infiltrative and infectious processes, metabolic and neuromuscular disorders, inherited syndromes and cardiomyopathies. These separate pathological mechanisms have the potential to lead to an elevated risk for adverse cardiac events which is not necessarily mitigated by the PPM implantation.^5-8^ Importantly, some of these disease processes may be reversible, omitting the need for permanent pacing. Several society guidelines have given a class IIa recommendation for performing advanced imaging as part of the pre-PPM implantation workup in patients who have conduction abnormalities and are suspected to have an underlying structural heart disease that was not apparent on echocardiography, particularly in those younger than 60 years.^9-11^ As information regarding the etiologies of “early-onset” complete atrioventricular block (CAVB) is scarce, we aimed to investigate potential causes for non-iatrogenic CAVB in younger patients and identify potential predictors for cardiac implantable electronic device (CIED) implantation in patients presenting with CAVB of unidentified etiology before 60 years of age.

## METHODS

### Data Source

The data were drawn from the National Inpatient Sample (NIS), the Healthcare Cost and Utilization Project (HCUP), and Agency for Healthcare Research and Quality (AHRQ).^12^ The NIS database includes only de-identified data; therefore, this study was deemed exempt from institutional review by the local Human Research Committee.

The NIS is the largest collection of all-payer data on inpatient hospitalizations in the United States. The dataset represents an approximate 20% stratified sample of all inpatient discharges from U.S. hospitals.^13^ This information includes patient-level and hospital-level factors such as patient demographic characteristics, primary and secondary diagnoses and procedures, co-morbidities, length of stay (LOS), hospital region, hospital teaching status, hospital bed size, and cost of hospitalization. National estimates can be calculated using the patient-level and hospital-level sampling weights that are provided by the HCUP.

For the purpose of this study, we obtained data from the years 2015 (last quarter) - 2019. International Classification of Diseases, 10^th^ Revision, Clinical Modification/Procedure Coding System (ICD-10-CM/PCS) was fully implemented from the last quarter of 2015 and thereafter for reporting diagnoses and procedures in the NIS database. For each index hospitalization, the database provides a principal discharge diagnosis and a maximum of 39 additional diagnoses, in addition to a maximum of 25 procedures.

### Study Population and Variables

We analyzed the NIS database from 2015 (last quarter) -2019 and identified all patients who had a primary diagnosis of CAVB using the ICD-10-CM code I442.Patients younger than 18 years and older than 60 years were excluded. We used ICD-10-CM/PCS codes (provided in Appendix Table 1) to identify and exclude patients who had any of the following diagnoses: mitral or aortic valve interventions, alcohol septal ablation or electrophysiological procedures that could raise suspicion of an iatrogenic cause for the conduction disorder. We also excluded patients with prior CIED implantations.

Patient level information including demographics, comorbidities, secondary diagnoses and clinically relevant procedures were collected from the database using ICD-10-CM/PCS codes (Appendix Table 1). Hospital level information and in-hospital outcomes including LOS and in-hospital mortality were analyzed from the database.

For the purposes of calculating Deyo-Charlson Comorbidity Index (Deyo-CCI), additional comorbidities were identified from the database using ICD-10-CM codes. Deyo-CCI is a modification of the Charlson Comorbidity Index, containing 19 comorbidity conditions with differential weights, with a total score ranging from 0 to 38 (Detailed information on Deyo-CCI provided in the Appendix Table 2).^14, 15^ Higher Deyo-CCI scores indicate a greater burden of comorbid diseases and are associated with mortality, one year after admission. The index has been used extensively in studies from administrative databases, with proved validity in predicting short- and long-term outcomes.^15-17^

Potential etiologies for CAVB, including, ischemic, infectious, inflammatory and infiltrative diseases, congenital heart disease, drug or metabolic/electrolyte disturbance induces were identified using ICD-10-CM codes (Appendix Table 1).

### Statistical Analysis

The Pearson’s chi-square (χ2) test and independent-samples t-test were used to compare categorical variables and continuous variables, respectively. The NIS provides discharge sample weights that are calculated within each sampling stratum as the ratio of discharges in the universe to discharges in the sample.^18^ We generated a weighted logistic regression models to identify independent predictors of CIED implantation in patients with CAVB of unidentified etiology. Candidate model variables included patient-level characteristics, Deyo-CCI, and hospital-level factors. We retained all predictor variables that were associated with in-hospital complications in our final multivariable regression models. For all analyses, we used SPSS^®^ software version 23 (IBM Corp., Armonk, NY). A p-value <0.05 was considered statistically significant.

## RESULTS

### Patient characteristics

We identified 2,899 patients aged 18-60 who were hospitalized with a primary diagnosis CAVB and without iatrogenic etiologies or in-situ CIED, meeting the inclusion and exclusion criteria for this study. By implementing the weighting method, this dataset represents an estimated total of 14,495 hospitalizations (Figure 1) for “early-onset” CAVB in the US, during the study period, average of about 4450 hospitalizations a year. The majority of patients (60%) were male, and the mean age of the cohort was 51.3 ± 9.1 years. Baseline characteristics of the study population are presented in detail in Table 1. Among the 14,495 patients, 3,050 (21%) had documentation of a prior conduction disorder, and 1,305 (9%) had a diagnosis of atrial fibrillation. 1,575 patients (11%) had a diagnosis of syncope, 930 (6%) had a diagnosis of cardiac arrest and 570 (4%) of cardiogenic shock.

**Figure 1:**
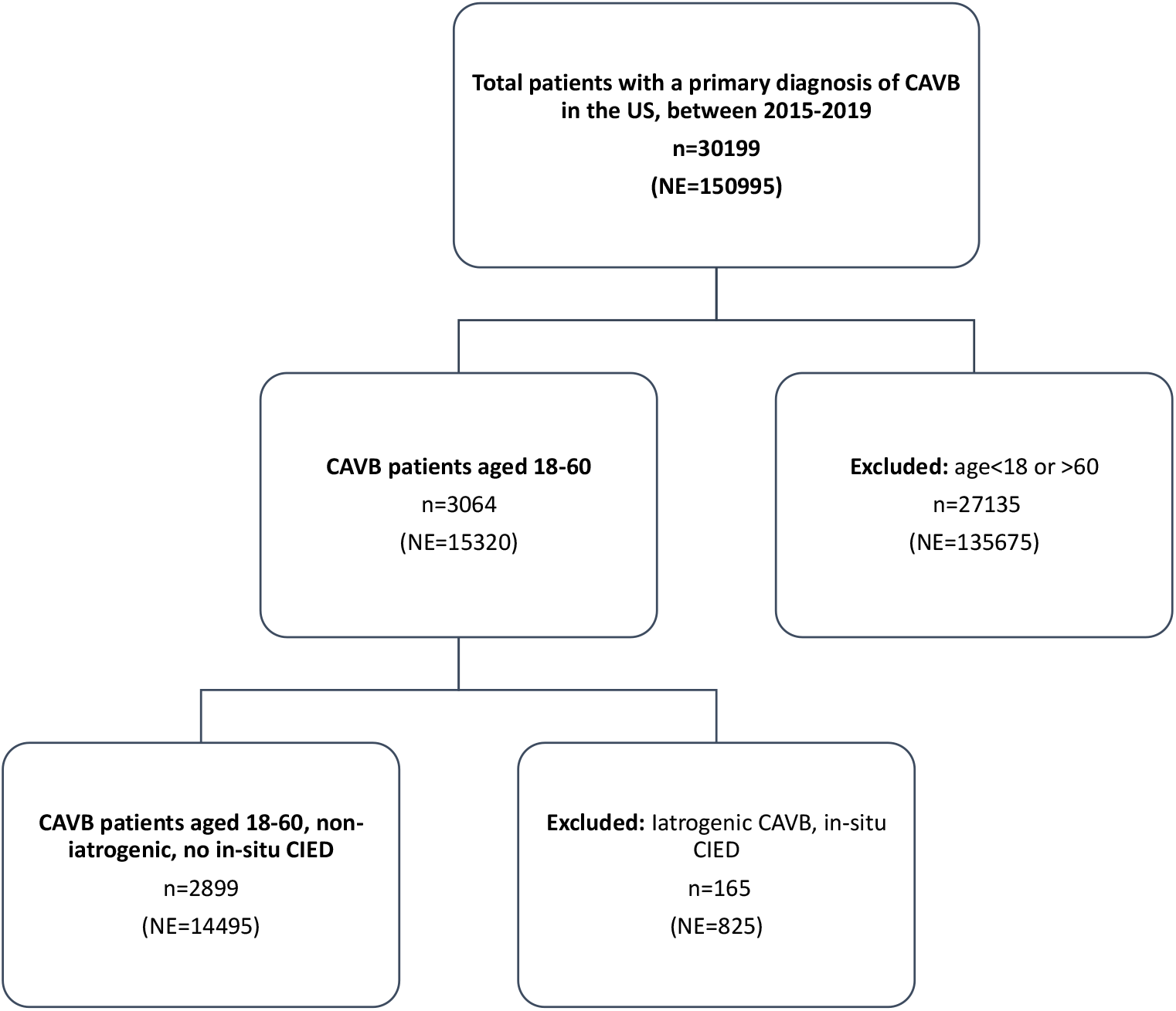
Flowchart of patient inclusion and exclusion. n= actual number of hospitalizations in the dataset, NE= national estimate of hospitalizations. National estimates of hospitalizations were calculated using sampling weights provided by the HCUP (see main text). CAVB= complete atrioventricular block, CIED= cardiac implantable electronic device, NE= national estimate.

**Table 1:**
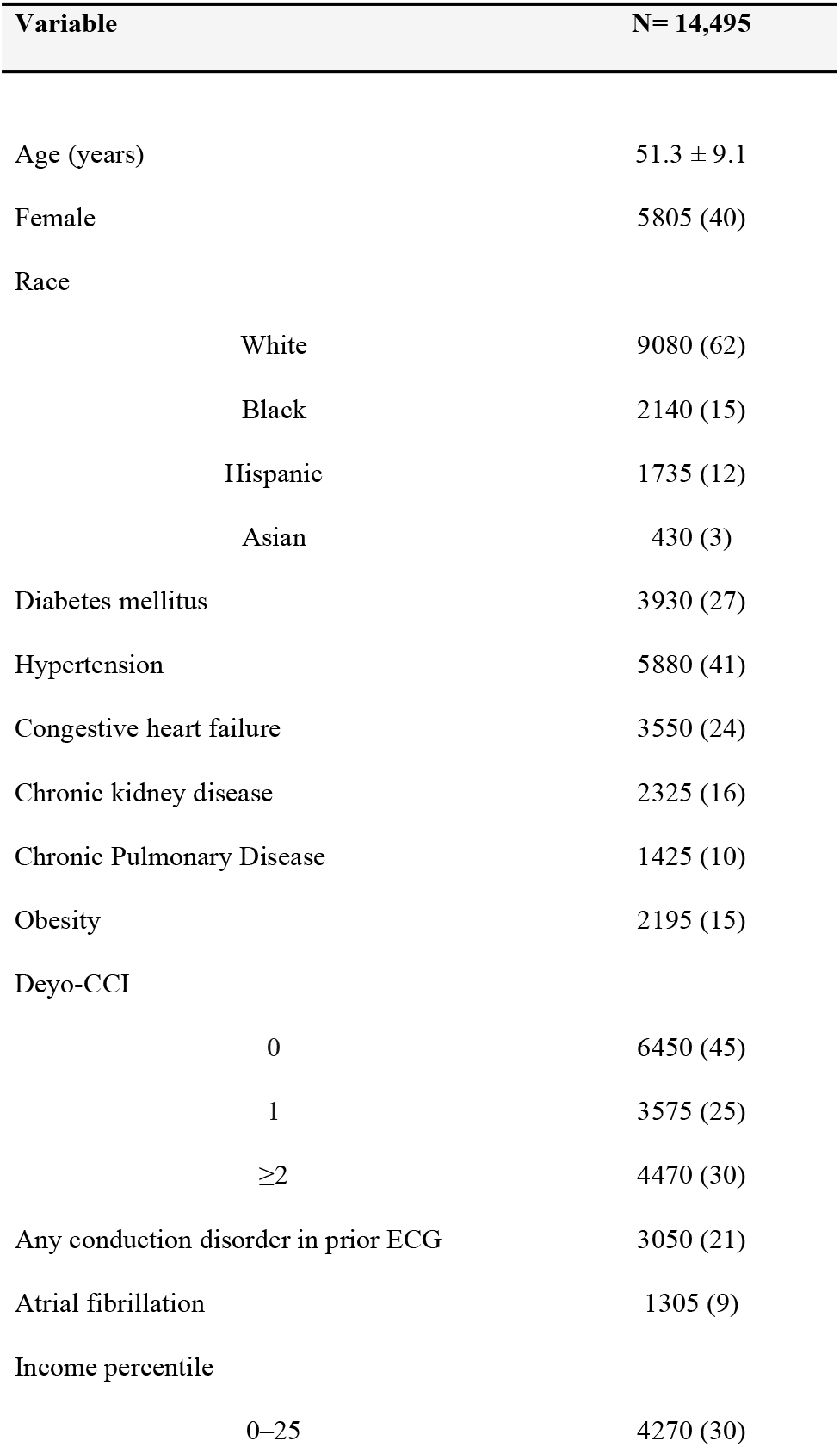
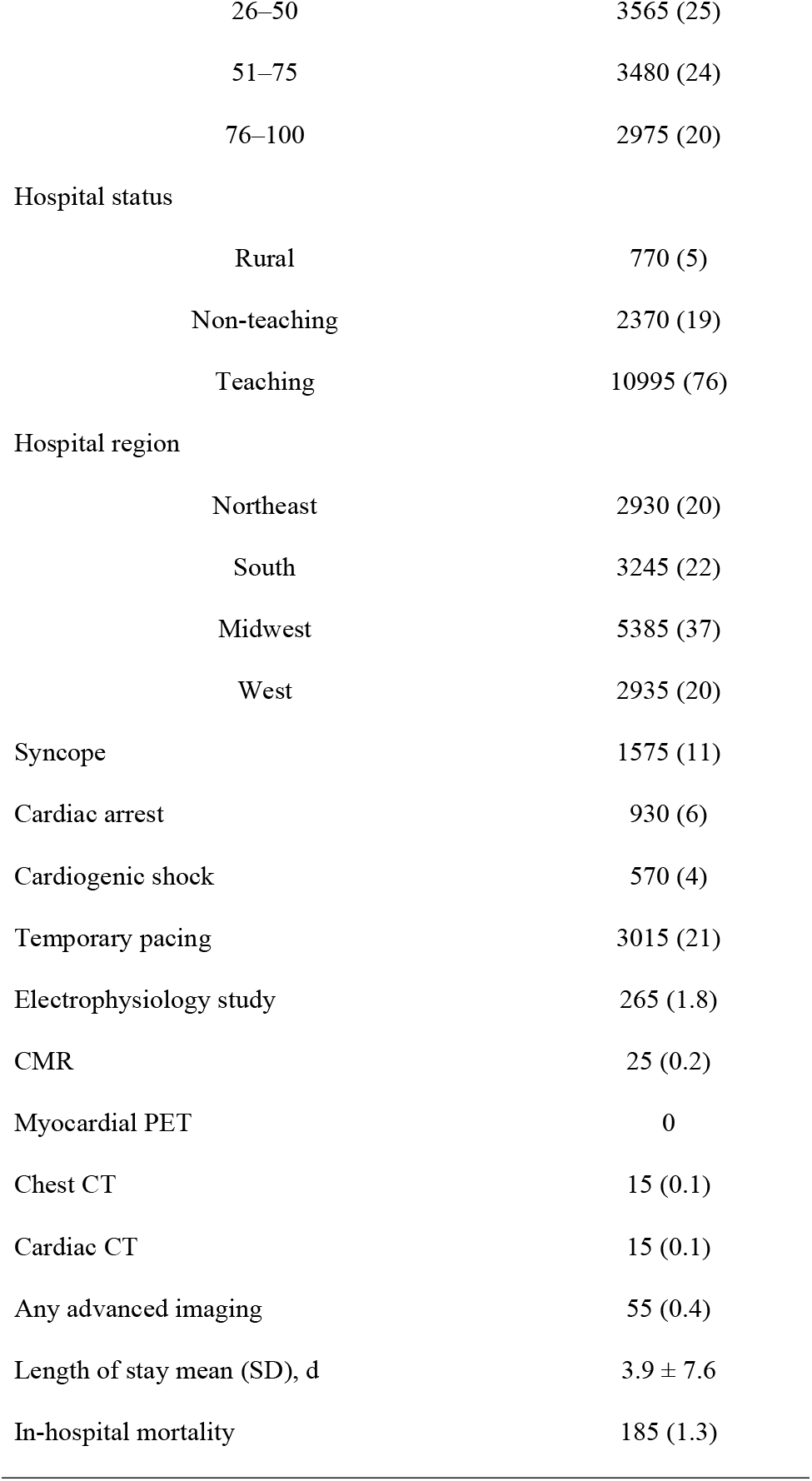
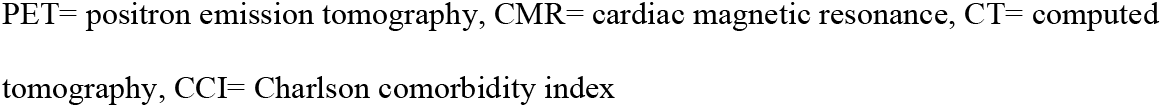
Demographics, risk factors and hospital characteristics. No. (%) of patients.

### Etiologies for non-iatrogenic CAVB

Potential etiologies for non-iatrogenic CAVB in patients aged 18-60 are presented in Table 2. The most common potentially reversible etiology was the identification of a metabolic disorder [n=1545 (10.9%)], among which hypoxia [n=935 (6.4%)] and acidosis [n=920 (6.3%)] were most common. Electrolyte imbalance was identified in 1,195 patients (8.2%) with hyperkalemia being the most common [n=1145 (7.9%)], while hypercalcemia and hypermagnesemia were identified in only 0.3% of patients.

**Table 2:** Etiologies for non-iatrogenic CAVB in patients aged 18-60. No. (%) of patients.

| Variable | N= 14495 |
| --- | --- |
| Unidentified etiology | 9735 (67.2) |
| Metabolic disorders | 1545 (10.9) |
| Non-Ischemic cardiomyopathy | 1540 (10.6) |
| Electrolyte imbalance | 1195 (8.2) |
| Myocardial infarction | 945 (6.8) |
| Ischemic cardiomyopathy | 450 (3.1) |
| Lyme disease | 260 (1.8) |
| Drug induced | 140 (1.0) |
| Congenital heart disease/AV block | 125 (0.9) |
| Cardiac sarcoidosis | 115 (0.8) |
| Endocarditis | 55 (0.4) |
| Myocarditis | 50 (0.3) |
| Muscular dystrophy | 35 (0.2) |
| Amyloidosis | 10 (0.1) |

A total of 1,540 patients (10.6%) had a diagnosis of non-ischemic cardiomyopathy (CM), of which 330 patients (2.3%) had documented dilated CM, 180 patients (1.2%) had hypertrophic CM, 55 patients (0.4%) had toxin mediated/alcoholic/restrictive CM and 985 patients (6.8%) had CM of unspecified etiology. There were 9,735 patients (67%) who had no clear potential etiology for CAVB identified.

### In-hospital course of patients with non-iatrogenic CAVB

Of the total population admitted for non-iatrogenic CAVB, 21% required temporary cardiac pacing. Interestingly, only a very small minority of patients had any advanced imaging work up during their index hospitalization for early-onset CAVB (0.2% CMR, 0% myocardial PET, 0.1% cardiac CT and 0.1% chest CT). A total of 265 patients (1.8%) underwent an electrophysiology study (EPS) during their hospitalization (Table 1).

The average LOS in the hospital for the study population was 3.9 ± 7.6 days. 185 patients (1.3%) died during their hospitalization.

### CIED implantation rates and predictors in patients with CAVB of unidentified etiology

Compared with patients with an identified etiology for CAVB (33% of 14495), which in some cases was reversible, patients with early-onset CAVB of unidentified etiology had a higher rate of any CIED implantation during their hospitalization (87% vs 74%, p<0.001), which was driven by a significantly higher rate of PPM implantations (84% vs 59%, p<0.001), while having a significantly lower rate of ICD (2% vs 15%, p<0.001) and CRT (3% vs 15%, p<0.001) implantations.

Among patients with early-onset CAVB of unidentified etiology, dichotomous age analysis revealed that PPM implantation rates were higher among patients over 40 years of age compared with those younger than 40 (87% vs 68%, p<0.001). No significant difference between patients older vs younger than 40 years was observed in either ICD (2% vs 2%, p=1.000) or CRT implantation rates (3% vs 2%, p=0.06) (Figure 2).

**Figure 2:**
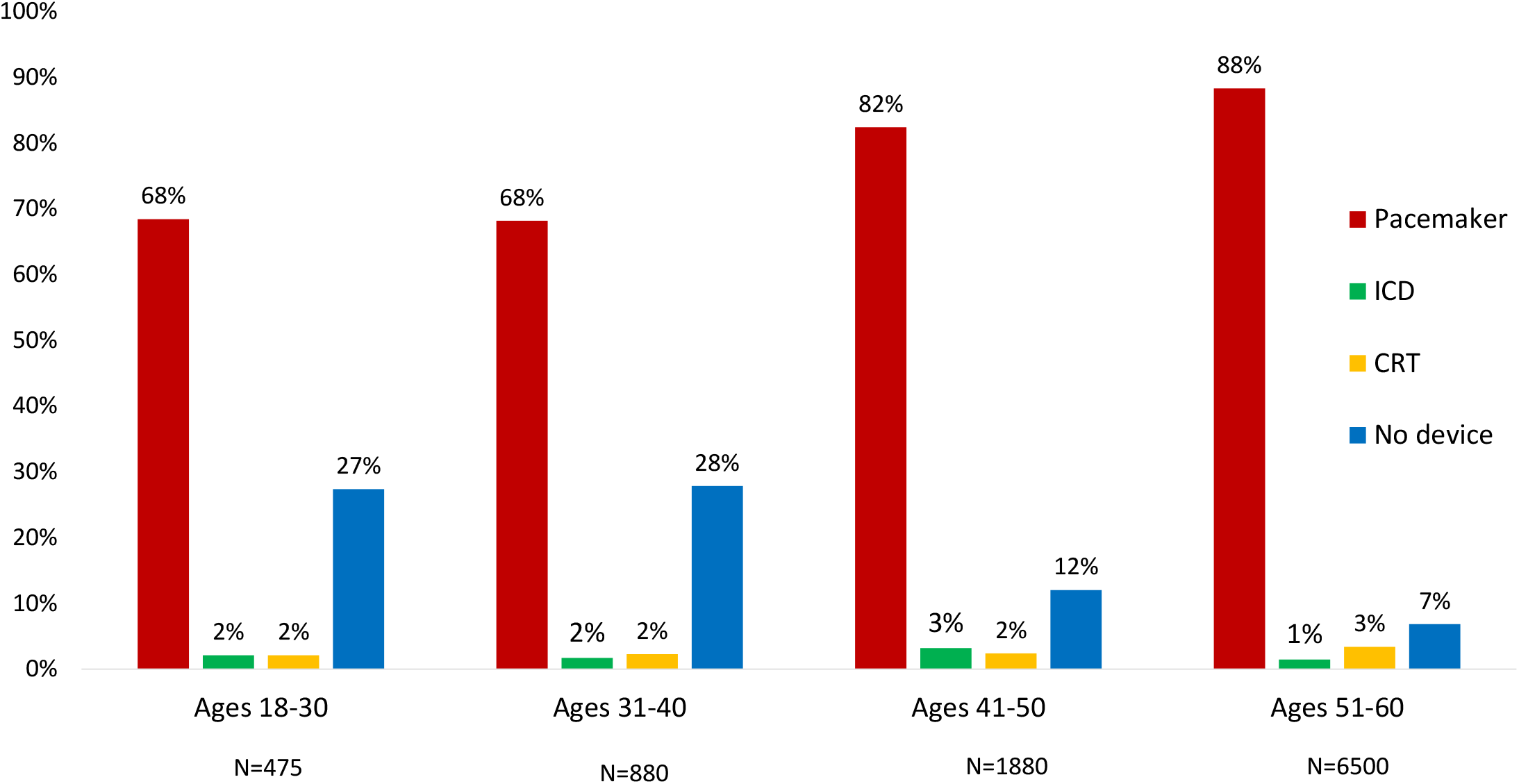
CIED implantation rates distribution by age groups in patients with CAVB of unidentified etiology. CIED= cardiac implantable electronic device, CRT= cardiac resynchronization therapy, ICD= implantable cardioverter defibrillator

Multivariate analyses adjusted for characteristics that were significantly associated with CIED implantation in patients with CAVB of unidentified etiology (Table 3), showed that a pre-existing atrioventricular conduction disorder was an independent predictor for implantation of CIED of any type [adjusted OR 1.51 (1.29-1.78), <0.001 for PPM; adjusted OR 2.70 (1.98-3.68), <0.001 for ICD; adjusted OR 2.03 (1.59-2.60), <0.001 for CRT]. Younger age was associated with ICD implantation [adjusted OR 0.98 (0.96-0.99), 0.01], while older age with PPM implantation [adjusted OR 1.05 (1.04-1.05), <0.001]. EPS performance was associated with ICD implantation [adjusted OR 3.26 (1.81-2.83), <0.001], and with a lower chance for PPM implantation [adjusted OR 0.41 (0.29-0.57), <0.001]. The need for TCP emerged as a predictor for PPM implantation [adjusted OR 1.54 (1.29-1.84), <0.001], while cardiac arrest was associated with ICD implantation [adjusted OR 2.78 (1.69-4.58), <0.001].

**Table 3:**
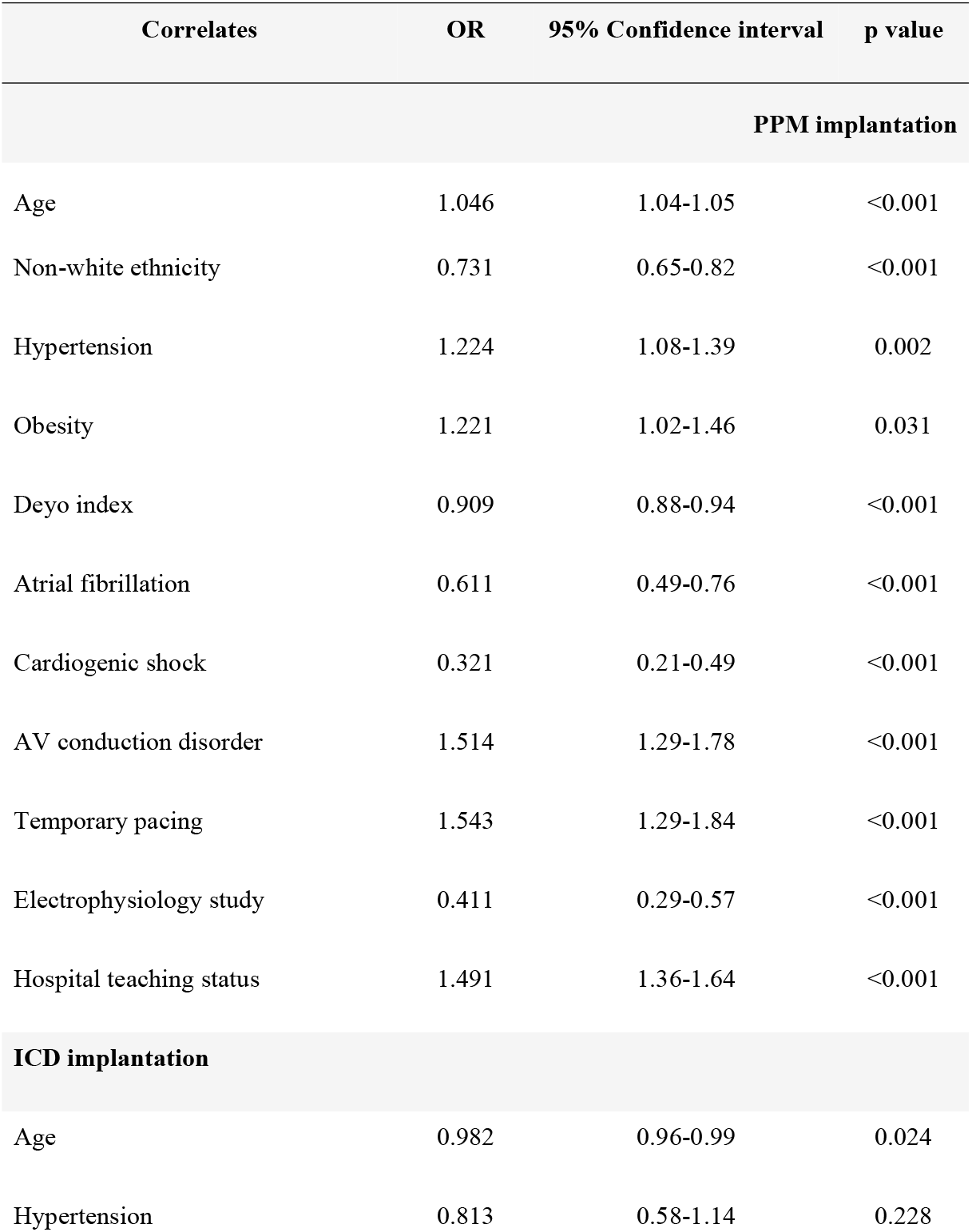
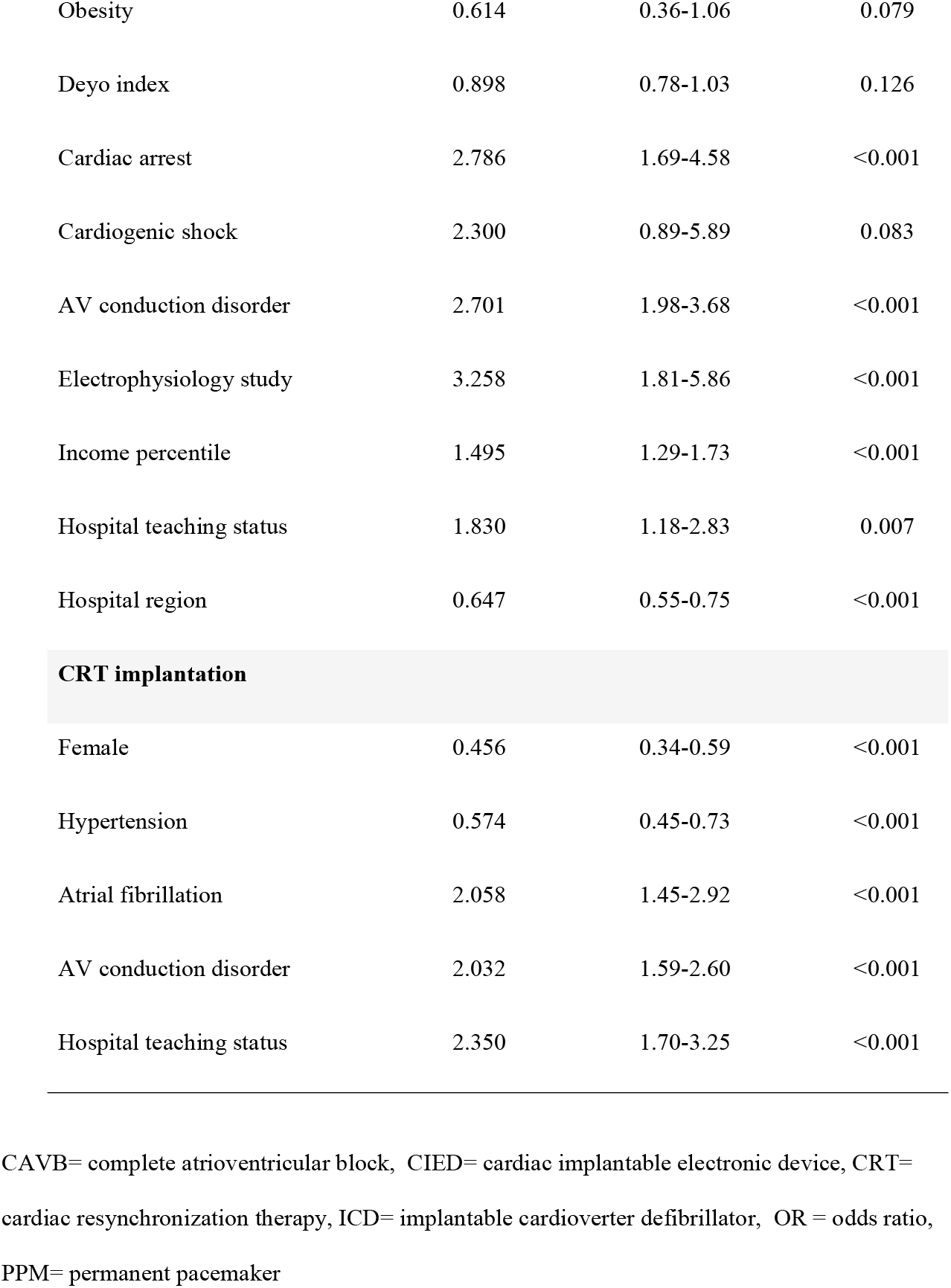
Predictors for CIED implantation in patients aged 18-60, with CAVB of unidentified etiology.

## DISCUSSION

Utilizing data from the NIS, the largest all-payer inpatient database in the U.S., we analyzed a weighted total of 14,495 hospitalizations for non-iatrogenic early-onset CAVB, between October 2015 and December 2019. Analysis of this nationwide real-world data revealed that 9,735 patients (67%) had no identifiable etiology for their conduction disorder. Among patients with early-onset CAVB of unidentified etiology, nearly 90% of patients were implanted with a CIED, which was in most cases a PPM. Importantly, despite society guidelines recommendations only 55 (0.4%) patients underwent any advanced imaging work up during their index hospitalization prior to the CIED placement.

CAVB is generally considered a medical emergency which prompts timely evaluation and management.^19^ The urgent workup performed in patients presenting with CAVB, especially if hemodynamically unstable, does not include the utilization of advanced cardiac imaging techniques. However, in our study only 10% of patients had an acute life-threatening condition (cardiac arrest or cardiogenic shock) and most patients (79%) did not require TCP. These data suggest that most patients in our study were potentially eligible for a more thorough evaluation for the etiology of their conduction disease prior to CIED implantation.

A myriad of disease processes, some of which are associated with adverse long-term prognosis, can affect the conduction system and may initially manifest solely as CAVB especially in younger patients. In studies that included patients aged<55-60 years who presented initially with unexplained advanced AVB, most patients had normal or near-normal left ventricular ejection fraction (LVEF) by echocardiography, 25%-34% of patients were diagnosed with cardiac sarcoidosis or giant cell myocarditis histologically or by advanced cardiac imaging.^7, 8^ In our study only 0.8% of patients had a diagnosis of cardiac sarcoidosis and 0.4% of myocarditis, and thus we cannot exclude that these conditions were underdiagnosed, in the general, nation-wide population, during the study period. Previous studies have shown that cardiac sarcoidosis, even if presenting initially as high degree AVB, is associated with a high risk for sudden cardiac death and ventricular tachycardia after PPM implantation.^20, 21^ Society guidelines emphasize the need for a more comprehensive pre-PPM implantation workup in young patients presenting with advanced AVB including performance of advanced cardiac imaging.^9-11^ If cardiac sarcoidosis is diagnosed in patients with an indication for permanent pacing, American guidelines and recent European guidelines give a class IIa recommendation for implanting an ICD.^9, 22, 23^ In our study only 2% of patients who had CAVB of unknown etiology received an ICD. Although younger age emerged as a significant predictor for ICD implantation in multivariable analysis, no significant difference was observed in ICD implantation rates between different age groups, and the majority of patients across all age groups who received a CIED were implanted with a PPM (Figure 2). Interestingly, EPS during hospitalization, which was performed in a small minority of patients, emerged as a predictor for ICD implantation or for avoidance of PPM placement (Table 3), possibly due to resulted arrhythmia induction or null results during EPS. These findings suggest that ICD therapy was potentially under-utilized in the US during our study period.

Very few studies evaluated the etiologies and prognosis of CAVB in younger patients.^5-8^ Rudbeck-Resdal et al conducted a study of 1,027 patients aged <50 years who were implanted with a PPM due to AVB in Denmark. In their report, 517 patients (50.3%) had no identifiable etiology. Interestingly, when patients with AVB secondary to cardiac surgery [n=157 (15.3%)], EP procedures [n=35 (3.4%)] and alcohol septal ablation [n=5 (0.5%)] complications were excluded from analysis, the rate of patients with CAVB of unknown etiology was 62%, akin to our results. The most common etiology of non-iatrogenic AVB in their report was congenital heart disease/AVB (13.2%), whereas in our study we observed a relatively low rate of these conditions (0.9%). This discrepancy can be explained by the inclusion of patients aged 0-18 years in their study, while we excluded pediatric patients in the current study.

In summary, while multiple etiologies can potentially underlie CAVB, especially in younger patients, most patients in our study were apparently managed in a “scoop and run” fashion, i.e. rapid, basic evaluation before proceeding to CIED implantation, which in most cases was a PPM. It was recently shown that young patients who were implanted with a PPM for AVB of unknown etiology, there was a higher rate of all-cause mortality and ventricular tachyarrhythmia compared to matched controls in long-term follow-up^5^ which emphasized the need for a more thorough pre-CIED implantation evaluation. However, as urgent management strategy is mandatory for some patients, post-PPM implantation surveillance and risk stratification is indicated. CMR has been shown to be safe in the presence of CIED^24^ and while image quality is dependent on the patient’s anatomy, device type and location, pacemakers perform better than ICD/CRTD systems in terms of artifacts and diagnostic yield.^25^

Several limitations need to be acknowledged. First, the NIS database is a retrospective administrative database that contains discharge-level records and as such is susceptible to coding errors. In addition, conditions with no specific ICD-CM-10 codes such as CAVB secondary to cardioinhibitory reflex or hereditary causes (e.g. LMNA and SCN5A mutations) could not be identified. It is conceivable that in a proportion of patients with CAVB of unidentified etiology, the conduction disorder was secondary to cardioinhibitory reflex, especially in younger patients who did not receive a CIED, or secondary to hereditary disorders in those who received a CIED during their hospitalization. Secondly, the lack of patient identifiers in the NIS database prevented us from identifying patients whose CIED implantation procedure was deferred for a later hospitalization or ambulatory visit, as only events that occurred in the index hospitalization were captured. The NIS database also does not include detailed information about patients’ clinical characteristics, medication, blood tests etc. Therefore, we cannot rule out residual confounding of the associations we observed. These limitations are counterbalanced by the real world, nationwide nature of the data, lack of selection bias as well as absence of reporting bias introduced by selective publication of results from specialized centers.

In conclusion, the majority patients hospitalized for non-iatrogenic early-onset CAVB had no identified etiology for the conduction disease during their index hospitalization, based on the nationwide NIS data. Despite the current guideline’s recommendation, advanced imaging prior to CIED implantation was under-utilized in this patient population. Nation-wide implementation of diagnostic and risk stratification algorithms is needed to improve adherence to guidelines, recommending the utilization of advanced imaging modalities in young patients with unexplained CAVB.

## Data Availability

The national database data used for this study, analytic methods, and study materials will not be made available to other researchers for purposes of reproducing the results or replicating the procedure because of restrictions on the sharing of data in the Healthcare Cost and Utilization Project Data Use Agreement. The NIS database is publicly available for purchase, and the transparent and detailed methods described in the manuscript make it possible for anyone who wishes to do so to replicate this study and reproduce our results

## Acknowledgments

None

## Figure legends

**Appendix Table 1:** ICD-10 CM/PCS codes used in data extraction.

| ICD-10 CM/PCS codes | Condition/Procedure |
| --- | --- |
| **Codes for potential iatrogenic causes for CAVB and for CIED in-situ** |  |
| 02RG07Z, 02RG08Z, 02RG0JZ, 02RG0KZ, 02RG37H,<br>02RG37Z, 02RG38H, 02RG38Z, 02RG3JH, 02RG3JZ,<br>02RG3KH, 02RG3KZ, 02RG47Z, 02RG48Z, 02RG4JZ,<br>02RG4KZ | Mitral valve interventions |
| 02RF07Z, 02RF08Z, 02RF0KZ, X2RF032, 02RF0JZ,<br>02RF37Z, 02RF38Z, 02RF3JZ, 02RF3KZ, X2RF332,<br>02RF37H, 02RF38H, 02RF3JH, 02RF3KH | Aortic valve interventions |
| 025M3ZZ | Alcohol septal ablation |
| 02563ZZ, 02573ZZ, 025K3ZZ, 025L3ZZ, 02583ZZ,<br>02553ZZ, 025M3ZZ, 025S3ZZ, 025T3ZZ | Electrophysiological procedures |
| Z950, Z95810 | CIED in-situ |
| **Codes for baseline characteristics and comorbidities not included in Deyo-CCI** |  |
| I10 | Hypertension |
| I48XX | Atrial fibrillation/flutter |
| E66X | Obesity |
| I440, I441, I447, I450, I451, I4510, I444, I445, I452, I453,<br>I454A | Any conduction disorder in prior ECG |
| **Codes for secondary hospitalization diagnoses** |  |
| R55, I459 | Syncope |
| I46X | Cardiac arrest |
| R570, R579 | Cardiogenic shock |
| **Codes for potential etiologies for CAVB** |  |
| I21X, I22X, I252 | Acute myocardial infarction |
| T461XXA, T447XX, T6589XA, T659XXA | Drug induced |
| E875, E8341, E8352 | Electrolyte imbalance |
| E035, J9601, J9611, J9621, J9691, E872, T68XXXX | Metabolic disorders |
| I255 | Ischemic cardiomyopathy |
| I42X | Non-ischemic cardiomyopathy |
| I330, I339 | Endocarditis |
| D8685 | Cardiac sarcoidosis |
| A692X | Lyme disease |
| I514 | Myocarditis |
| E854 | Cardiac amyloidosis |
| Q205, Q213, Q210, Q24X | Congenital heart disease/AV block |
| G710, G7100 | Muscular dystrophy |
| **Codes for diagnostic and interventional procedures during hospitalization** |  |
| B236Y0Z, B236YZZ, B236ZZZ, B231Y0Z, B231YZZ, B231ZZZ, B233Y0Z, B233YZZ, B233ZZZ, B231Y0Z, B231YZZ, B231ZZZ, B233Y0Z, B233YZZ, B233ZZZ, B236Y0Z, B236YZZ, B236ZZZ | Cardiac magnetic resonance |
| C23GKZZ, C23GYZZ | Myocardial positron emission tomography |
| BB2400Z, BB240ZZ, BB2410Z, BB241ZZ, BB24Y0Z, BB24YZZ, BB24ZZZ, BW241ZZ, BW241ZZ, | Chest computed tomography |
| BW24YZZ, BW24ZZZ, BW25ZZZ, BW250ZZ,<br>BW251ZZ, BW25YZZ |  |
| B22600Z, B2260ZZ, B22610Z, B2261ZZ, B226Y0Z,<br>B226YZZ, B226Z2Z, B226ZZZ, B22100Z, B2210ZZ,<br>B22110Z, B2211ZZ, B221Y0Z, B221YZZ, B221Z2Z,<br>B221ZZZ, B22300Z, B2230ZZ, B22310Z, B2231ZZ,<br>B223Y0Z, B223YZZ, B223Z2Z, B223ZZZ | Cardiac computed tomography |
| 4A0234Z, 4A023PZ, 4A023FZ, 02K80ZZ, 02K83ZZ,<br>02K84ZZ | Electrophysiology study |
| 5A12X3Z | Temporary cardiac pacing |
| 02HK3JZ, 02HK4JZ, 0JH605Z, 0JH635Z, 0JH606Z,<br>0JH636Z, 02H64JZ, 02H63JZ, 0JH604Z, 0JH634Z,<br>0JH804Z, 0JH834Z, 0JH805Z, 0JH835Z, 0JH806Z,<br>0JH836Z | Permanent pacemaker implantation |
| 02HK3KZ, 02HK4KZ, 0JH608Z, 0JH638Z, 0JH808Z,<br>0JH838Z | Implantable cardioverter-defibrillator<br>implantation |
| 0JH607Z, 0JH637Z, 0JH609Z, 0JH639Z, 02H43JZ,<br>02H44JZ, 02H40JZ, 02H40NZ, 02H43NZ, 02H44NZ,<br>0JH807Z, 0JH837Z, 0JH809Z, 0JH839Z | Cardiac resynchronization therapy |

**Appendix Table 2:** ICD-10 CM Codes for conditions incorporated in Deyo-CCI, and scoring system used to compute CCI scores.

| ICD-10 CM codes | Condition | Score |
| --- | --- | --- |
| I21.x, I22.x, I25.2 | Myocardial infarction | 1 |
| I11.0, I13.0, I13.2, I25.5, I42.0, I42.5-I42.9, I43.x, I50.x, P29.0 | Congestive heart failure | 1 |
| I70.x, I71.x, I73.1, I73.8, I73.9, I77.1, I79.0, I79.1, I79.8, K55.1, K55.8, K55.9, Z95.8, Z95.9 | Peripheral vascular disease | 1 |
| G45.x, G46.x, H34.0x, H34.1x, H34.2x, I60.x-I68.x | Cerebrovascular disease | 1 |
| F01.x-F03.x, F04, F05, F06.1, F06.8, G13.2, G13.8, G30.x, G31.0x, G31.1, G31.2, G91.4, G94, R41.81, R54 | Dementia | 1 |
| J40.x-J47.x, J60.x-J67.x, J68.4, J70.1, J70.3 | Chronic pulmonary disease | 1 |
| M05.x, M06.x, M31.5, M32.x-M34.x, M35.1, M35.3, M36.0 | Rheumatologic disease | 1 |
| K25.x-K28.x | Peptic ulcer disease | 1 |
| B18.x, K70.0-K70.3, K70.9, K71.3-K71.5, K71.7, K73.x, K74.x, K76.0, K76.2-K76.4, K76.8, K76.9, Z94.4 | Mild liver disease | 1 |
| Main codes: E08, E09, E10, E11, E13.<br>Relevant subcodes: E**.0x, E**.1x, E**.6x, E**.8x, E**.9x | Diabetes without chronic complications | 1 |
| I12.9, I13.0, I13.10, N03.x, N05.x, N18.1-N18.4, N18.9, Z94.0 | Renal disease, mild or moderate | 1 |
| Main codes: E08, E09, E10, E11, E13. | Diabetes with chronic complications | 2 |
| Relevant subcodes: E**.2x, E**.3x, E**.4x, E**.5x |  |  |
| G04.1, G11.4, G80.0, G80.1, G80.2, G81.x, G82.x, G83.x | Hemiplegia or paraplegia | 2 |
| C0x.x, C1x.x, C2x.x, C30.x-C34.x, C37.x-C41.x, C43.x, C45.x-C58.x, C60.x-C63.x, C76.x, C80.1, C81.x-C85.x, C88.x, C9x.x | Any malignancy including leukemia and lymphoma except malignant nonmelanoma of the skin | 2 |
| I85.0x, I86.4, K70.4x, K71.1x, K72.1x, K72.9x, K76.5, K76.6, K76.7 | Moderate or severe liver disease | 3 |
| I12.0, I13.11, I13.2, N18.5, N18.6, N19.x, N25.0, Z49.x, Z99.2 | Renal disease, severe | 3 |
| B20.x | HIV infection | 3 |
| C77.x-C79.x, C80.0, C80.2 | Metastatic solid tumor | 6 |
| B37.x, C53.x, B38.x, B45.x, A07.2, B25.x, G93.4x, B00, B39.x, A07.3, C46.x, C81-96, A31.x, A15-19, B59, Z87.01, A81.2, A02.1, B58.x, R64 | Acquired Immunodeficiency syndrome (AIDS) | 6 |

## References

1. Friedberg CK, Donoso E, Stein WG. Nonsurgical Acquired Heart Block. Ann N Y Acad Sci. 1964;111:835–847.

2. Udo EO, van Hemel NM, Zuithoff NP, Doevendans PA, Moons KG. Prognosis of the bradycardia pacemaker recipient assessed at first implantation: a nationwide cohort study. Heart. 2013;99(21):1573–1578.

3. Lev M. Anatomic Basis for Atrioventricular Block. Am J Med. 1964;37:742–748.

4. Lenegre J. Etiology and Pathology of Bilateral Bundle Branch Block in Relation to Complete Heart Block. Prog Cardiovasc Dis. 1964;6:409–444.

5. Dideriksen JR, Christiansen MK, Johansen JB, Nielsen JC, Bundgaard H, Jensen HK. Long-term outcomes in young patients with atrioventricular block of unknown aetiology. Eur Heart J. 2021;42(21):2060–2068.

6. Rudbeck-Resdal J, Christiansen MK, Johansen JB, Nielsen JC, Bundgaard H, Jensen HK. Aetiologies and temporal trends of atrioventricular block in young patients: a 20-year nationwide study. Europace. 2019;21(11):1710–1716.

7. Kandolin R, Lehtonen J, Kupari M. Cardiac sarcoidosis and giant cell myocarditis as causes of atrioventricular block in young and middle-aged adults. Circ Arrhythm Electrophysiol. 2011;4(3):303–309.

8. Nery PB, Beanlands RS, Nair GM, et al. Atrioventricular block as the initial manifestation of cardiac sarcoidosis in middle-aged adults. J Cardiovasc Electrophysiol. 2014;25(8):875–881.

9. Birnie DH, Sauer WH, Bogun F, et al. HRS expert consensus statement on the diagnosis and management of arrhythmias associated with cardiac sarcoidosis. Heart Rhythm. 2014;11(7):1305–1323.

10. Glikson M, Nielsen JC, Kronborg MB, et al. 2021 ESC Guidelines on cardiac pacing and cardiac resynchronization therapy. Eur Heart J. 2021;42(35):3427–3520.

11. Kusumoto FM, Schoenfeld MH, Barrett C, et al. 2018 ACC/AHA/HRS Guideline on the Evaluation and Management of Patients With Bradycardia and Cardiac Conduction Delay: A Report of the American College of Cardiology/American Heart Association Task Force on Clinical Practice Guidelines and the Heart Rhythm Society. Circulation. 2019;140(8):e382–e482.

12. Introduction to the HCUP National Inpatient Sample (NIS). The National (nationwide) Inpatient Sample database documentation. 2020. https://www.hcup-us.ahrq.gov/overview.jsp

13. Steiner C, Elixhauser A, Schnaier J. The healthcare cost and utilization project: an overview. Eff Clin Pract. 2002;5(3):143–151.

14. Deyo RA, Cherkin DC, Ciol MA. Adapting a clinical comorbidity index for use with ICD-9-CM administrative databases. J Clin Epidemiol. 1992;45(6):613–619.

15. Glasheen WP, Cordier T, Gumpina R, Haugh G, Davis J, Renda A. Charlson Comorbidity Index: ICD-9 Update and ICD-10 Translation. Am Health Drug Benefits. 2019;12(4):188–197.

16. Chu YT, Ng YY, Wu SC. Comparison of different comorbidity measures for use with administrative data in predicting short- and long-term mortality. BMC Health Serv Res. 2010;10:140.

17. Radovanovic D, Seifert B, Urban P, et al. Validity of Charlson Comorbidity Index in patients hospitalised with acute coronary syndrome. Insights from the nationwide AMIS Plus registry 2002-2012. Heart. 2014;100(4):288–294.

18. Hosseini SM, Moazzami K, Rozen G, et al. Utilization and in-hospital complications of cardiac resynchronization therapy: trends in the United States from 2003 to 2013. Eur Heart J. 2017;38(27):2122–2128.

19. Panchal AR, Bartos JA, Cabanas JG, et al. Part 3: Adult Basic and Advanced Life Support: 2020 American Heart Association Guidelines for Cardiopulmonary Resuscitation and Emergency Cardiovascular Care. Circulation. 2020;142(16_suppl_2):S366–S468.

20. Takaya Y, Kusano KF, Nakamura K, Ito H. Outcomes in patients with high-degree atrioventricular block as the initial manifestation of cardiac sarcoidosis. Am J Cardiol. 2015;115(4):505–509.

21. Nordenswan HK, Lehtonen J, Ekstrom K, et al. Outcome of Cardiac Sarcoidosis Presenting With High-Grade Atrioventricular Block. Circ Arrhythm Electrophysiol. 2018;11(8):e006145.

22. Al-Khatib SM, Stevenson WG, Ackerman MJ, et al. 2017 AHA/ACC/HRS Guideline for Management of Patients With Ventricular Arrhythmias and the Prevention of Sudden Cardiac Death: A Report of the American College of Cardiology/American Heart Association Task Force on Clinical Practice Guidelines and the Heart Rhythm Society. Circulation. 2018;138(13):e272–e391.

23. Zeppenfeld K, Tfelt-Hansen J, de Riva M, et al. 2022 ESC Guidelines for the management of patients with ventricular arrhythmias and the prevention of sudden cardiac death. Eur Heart J. 2022.

24. Roguin A, Zviman MM, Meininger GR, et al. Modern pacemaker and implantable cardioverter/defibrillator systems can be magnetic resonance imaging safe: in vitro and in vivo assessment of safety and function at 1.5 T. Circulation. 2004;110(5):475–482.

25. Hilbert S, Jahnke C, Loebe S, et al. Cardiovascular magnetic resonance imaging in patients with cardiac implantable electronic devices: a device-dependent imaging strategy for improved image quality. Eur Heart J Cardiovasc Imaging. 2018;19(9):1051–1061.

